# A Novel Approach to Monitoring the COVID-19 Pandemic using Emergency Department Discharge Diagnoses

**DOI:** 10.1101/2020.06.09.20126508

**Authors:** Herbert C. Duber, M. Kennedy Hall, Karl Jablonowski, Susan A. Stern, Ali H Mokdad, Abraham D Flaxman

## Abstract

**Introduction:** Tracking the COVID-19 pandemic using existing metrics such as confirmed cases and deaths are insufficient for understanding the trajectory of the pandemic and identifying the next wave of cases. In this study, we demonstrate the utility of monitoring the daily number of patients with COVID-like illness (CLI) who present to the Emergency Department (ED) as a tool that can guide local response efforts.

**Methods:** Using data from two hospitals in King County, WA, we examined the daily volume of CLI visits, and compare them to confirmed COVID cases and COVID deaths in the County. A linear regression model with varying lags is used to predict the number of daily COVID deaths from the number of CLI visits.

**Results:** CLI visits appear to rise and peak well in advance of both confirmed COVID cases and deaths in King County. Our regression analysis to predict daily deaths with a lagged count of CLI visits in the ED showed that the R^2^ value was maximized at 14 days.

**Conclusions:** ED CLI visits are a leading indicator of the pandemic. Adopting and scaling up a CLI monitoring approach at the local level will provide needed actionable evidence to policy makers and health officials struggling to confront this health challenge.

## Introduction

Current data suggests that the US has passed the peak of the initial COVID-19 outbreak. However, as social distancing measures are eased, there is significant concern for a resurgence.^1^ Unfortunately, the most commonly used metrics for assessing daily progress – case counts and deaths – have significant limitations.^2^ Case counts are highly uncertain due to asymptomatic carriage, limited testing capacity, and an uneven distribution of testing across populations.^3^ Deaths may occur weeks after symptom onset and still requires definitive diagnosis.^4^ In this study, we demonstrate the utility of monitoring the daily number of patients with COVID-like illness (CLI) who present to the Emergency Department (ED) as a tool that addresses these limitations and can guide local response efforts.

## Methods

We obtained de-identified ED visit data from two hospitals (Harborview and University of Washington Medical Centers) located in Seattle, King County, WA. Daily confirmed COVID-19 case and death counts in King County are publically available and published online (https://github.com/nytimes/covid-19-data).

Prior work has demonstrated that hospitalized patients with SARS-CoV-2 usually present with influenza-like illness (ILI) symptoms,^5^ and co-infection with respiratory syncytial virus (RSV) and influenza is uncommon. ^6^ Therefore, we defined ED visits as CLI if they tested positive for COVID or they met the Armed Forces Health Surveillance Center (AFHSC) ICD-10 case definition for ILI (https://health.mil/Reference-Center/Publications/2015/10/01/Influenza-Like-Illness), and did not test positive for either influenza or RSV. In order to account for other viral pathogens, we established a baseline of visits that met CLI criteria in the pre-COVID era (January 18-31, 2020). The mean number of visits per day meeting this definition during the pre-COVID era was then subtracted from daily CLI visit counts in the COVID-era beginning March 1, 2020 to identify the number of daily CLI visits.

We examined visually and quantitatively the daily volume of CLI visits, confirmed COVID cases and COVID deaths. Because all of these variables are noisy, we smoothed them with a seven-day triangular window. To determine the degree to which CLI visits lead COVID deaths, we used a linear regression model to predict the number of daily COVID deaths on day *t* from the number of CLI visits on day *t− l* for a range of lags *l*. Since it is illogical to predict more than zero COVID deaths on the basis of zero CLI visits, we fixed the intercept to be zero, and used the regression equation death s_*t*_∼ *β* ·CLI_*t−l*_+*∈*_*t*_. For each lag *l*, we calculated the *R*^2^value as a goodness-of-fit metric, and found the lag that maximized *R*^2^.

## Results

A total of 27,288 ED visits were recorded between January 1 and May 1, 2020. 422 visits were classified as CLI. 29.3% of CLI visits resulted in admission. CLI visits appear to rise and peak well in advance of both confirmed COVID cases and deaths in King County (Figure 1). The smoothed peak of CLI visits occurs on March 25, whereas confirmed COVID cases and deaths peak on April 8 and April 9, respectively.

**Fig 1:**
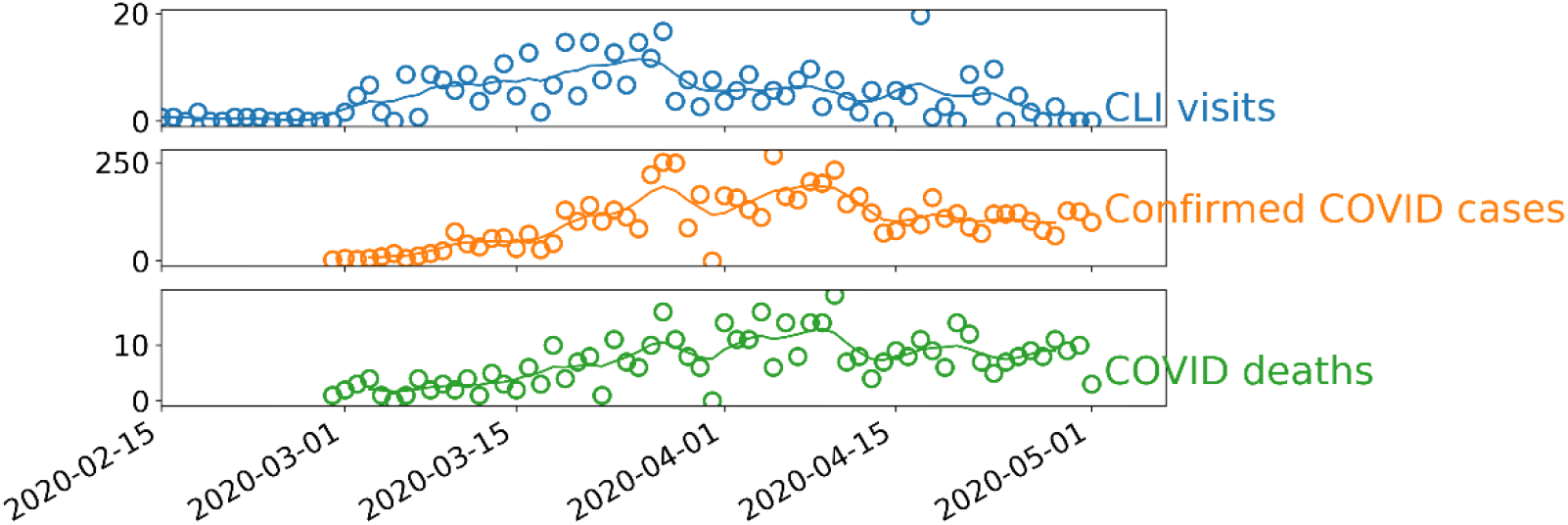
Daily count and smoothed time series of CLI visits, confirmed COVID-19 cases and COVID-19 deaths.

Our regression analysis to predict daily deaths with a lagged count of CLI visits in the ED showed that the R^2^ value was maximized by predicting the number of daily deaths that would occur 14 days after CLI visits (Figure 2).

**Fig 2:**
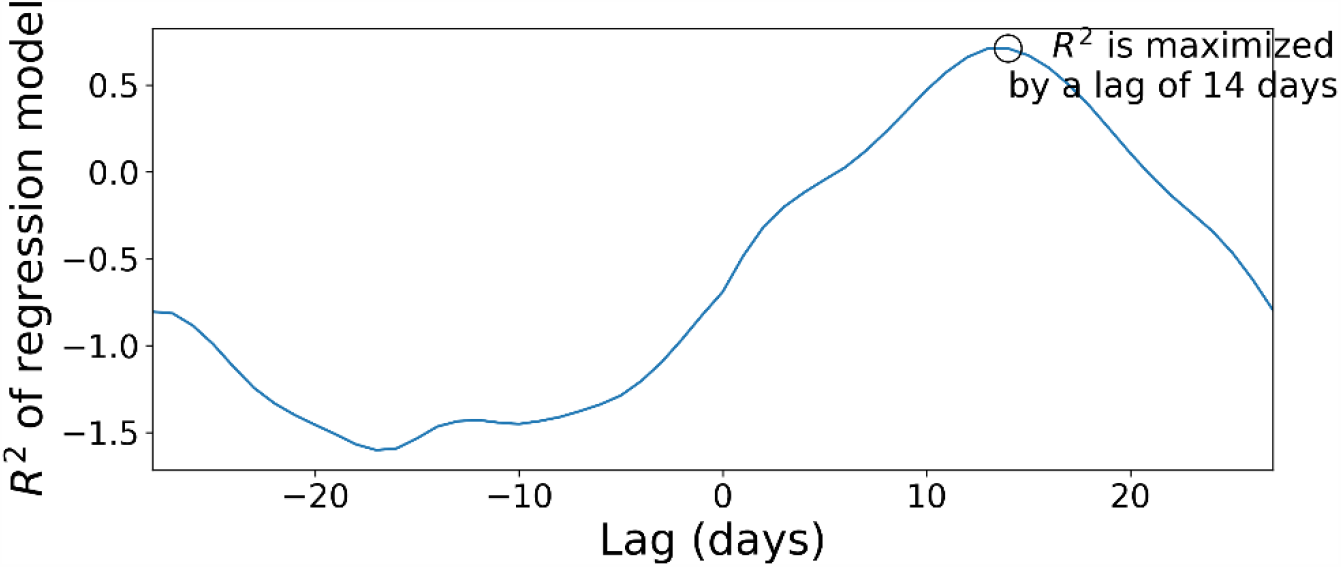
Relationship between CLI visits and COVID-19 deaths. Varying lag on the X axis and *R*^2^ on the Y axis. *R*^2^ is maximized using a lag of 14 days.

## Discussion

In this study, we demonstrated that ED-based CLI visits were a leading indicator of the COVID-19 pandemic in King County, WA. Moreover, our observation of declining CLI visits is consistent with local trends showing a declining number of admitted COVID-19 patients in our hospitals and a county-wide slowing of deaths. Therefore, we propose local CLI monitoring as an innovative, simple and timely approach to monitor COVID-19 in the US and elsewhere.

As we enter into a period of sustained low-level community transmission, and a national strategy based on differentiated policy at the state and local level, early identification of hotspots is critical. While some locations may have the capacity to test every individual for COVID-19, many do not. Further, while admitted COVID-19 cases can be used in a similar fashion, it still requires confirmatory testing and draws from a smaller sample, especially in locations with smaller populations and low overall COVID-19 prevalence. Adopting and scaling up a CLI monitoring approach at the local level will provide needed actionable evidence to policy makers and health officials struggling to confront this health challenge.

## Data Availability

COVID-19 case and death counts are available at the website provided. Hospital data is unavailable.

https://github.com/nytimes/covid-19-data

